# Analysis of the prediction of the 2021 time-evolution of the Covid-19 Pandemic in Italy using a Planck’s distribution

**DOI:** 10.1101/2021.09.06.21263173

**Authors:** Ignazio Ciufolini, Antonio Paolozzi

**Affiliations:** Dipartimento di Ingegneria dell’Innovazione, University of Salento, Lecce and Centro Fermi, Rome, Italy; Scuola di Ingegneria Aerospaziale, Sapienza, University of Rome

## Abstract

In a previous paper we studied the time evolution of the Covid-19 pandemic in Italy during the first wave of 2020 using a number of distribution laws. We concluded that the best distribution law to predict the evolution of the pandemic, if basic conditions of the pandemic (such as distancing measures, use of masks, start of schools, intensive use of public transportation, beginning and end of holidays, vaccination campaign and no significant onset of new Covid variants) do not appreciably change, is a distribution of the type of Planck’s law with three parameters. In our 2020 study we did not use the number of daily positive cases in Italy but the ratio of daily positive cases per number of daily tests, ratio today sometimes referred to as: “*positivity rate*”. We showed that, if basic conditions do not change, the Planck’s distribution with three parameters provides very good predictions of the *positivity rate* about one month in advance. In particular, in a second paper, using the Planck’s distribution with three parameters, we predicted, about one month in advance, the spread of the pandemic in Italy during the Christmas 2020 holidays with an error of a few percent only. We then study the present (September 2021) evolution of the pandemic in Italy and we show that the Planck’s distribution, based on the data of July and August, predicts well the evolution of the pandemic. In particular, we show that the peak of the *positivity rate* was predicted to occur approximately around the middle of August and that the agreement of this Planck’s function (obtained fitting the data up to 10 July 2021) and the *positivity rate* observed after 5 weeks, on 12 September 2021 is very good. However, the end of the Italian holidays and the start of all the activities including schools, intensive use of public transportation and further changes in distancing measures may cause a discrepancy of the predicted trend of the *positivity rate* of the pandemic with respect to the real observed values.

## 1. Discussion of previous analysis

In a previous paper [1], in the attempt to mathematically predict the evolution of the pandemic in Italy, we fitted the ratio of daily cases per daily tests, or “*positivity rate*”, from February 26, 2020 to the beginning of April 2020, using a number of different distributions (the number of daily cases and daily tests were taken from [2]). We considered distributions of the type of Gauss, Beta, Gamma, Weibull, Lognormal and two ones of the type of the Planck’s black body radiation law. The number of independent parameters chosen for some distributions was between two and three. It turned out that a distribution of the type of the Planck’s law with three independent parameters, to be experimentally fitted, provided the best prediction capability up to the end of April 2020, i.e., about one month in advance. The Planck-type distribution law we used is:

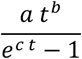

Where a, b and c are three parameters to be fitted for. Planck’s law is one of the outstanding laws of physics [3], it describes the energy of the radiation, emitted by a black body at thermal equilibrium with temperature *T*, as a function of the frequency or of the wavelength. For example, the number of photons emitted per unit area, unit time and unit frequency *ν*, is:

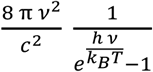

where *h, c* and *k*_*B*_ are Planck constant, speed of light and Boltzmann constant respectively. Derived in 1900 by Max Planck, with the idea of energy quantization, marked the beginning of Quantum Mechanics. A distribution of the type of the Planck’s law with three parameters showed a good prediction potentiality over a period of about one month also in the second wave of Covid 19.

Indeed, in [4], using 36 days starting from 17 October 2020 up to 21 November 2020 (included), we predicted, at the end of November 2020, the behavior of the second wave of the pandemic in Italy using the Planck’s distribution with three parameters. Fig. 1 is the prediction (corresponding to Fig. 4 published in [4]), of the daily cases-to-test ratios (*positivity rate*). The continuous curve is the best fit of the 36 data using a three parameter Planck’s law. Christmas 2020 is marked with the vertical blue line. This analysis [4] published on 9 December 2020 in the medRxiv, predicted quite well the behavior of the pandemic in Italy at the Christmas 2020 holidays (see Fig. 2), i.e., after approximately one month from the last experimental data of 21 November 2020 (included). For example, after 31 days, on 22 December 2020, the difference between our prediction and the observed positivity rate has shown an error of less than 2 percent only. Figure 2 shows our prediction (solid curve), based on the fit of the circles in red (positivity rates of 36 days from 17 October 2020, included, until 21 November 2020) with the Planck’s distribution, together with the positivity rates later observed (which were not known to us at the end of November 2020) which are shown by the black circles. The percent error of less than 2 percent only in the prediction of the spread of the pandemic is calculated (Fig. 2) as difference between the solid curve and the real observed cases on 22 December 2020. Starting approximately from 25 December 2020, there was a small peak in the positivity rates, probably due to the attenuation of social distancing during the Christmas holiday season. However, after such small peak lasting about three weeks from Christmas, the Planck’s curve is still fitting quite well the subsequent positivity rates up to, e.g., 19 February 2021.

**Fig. 1.**
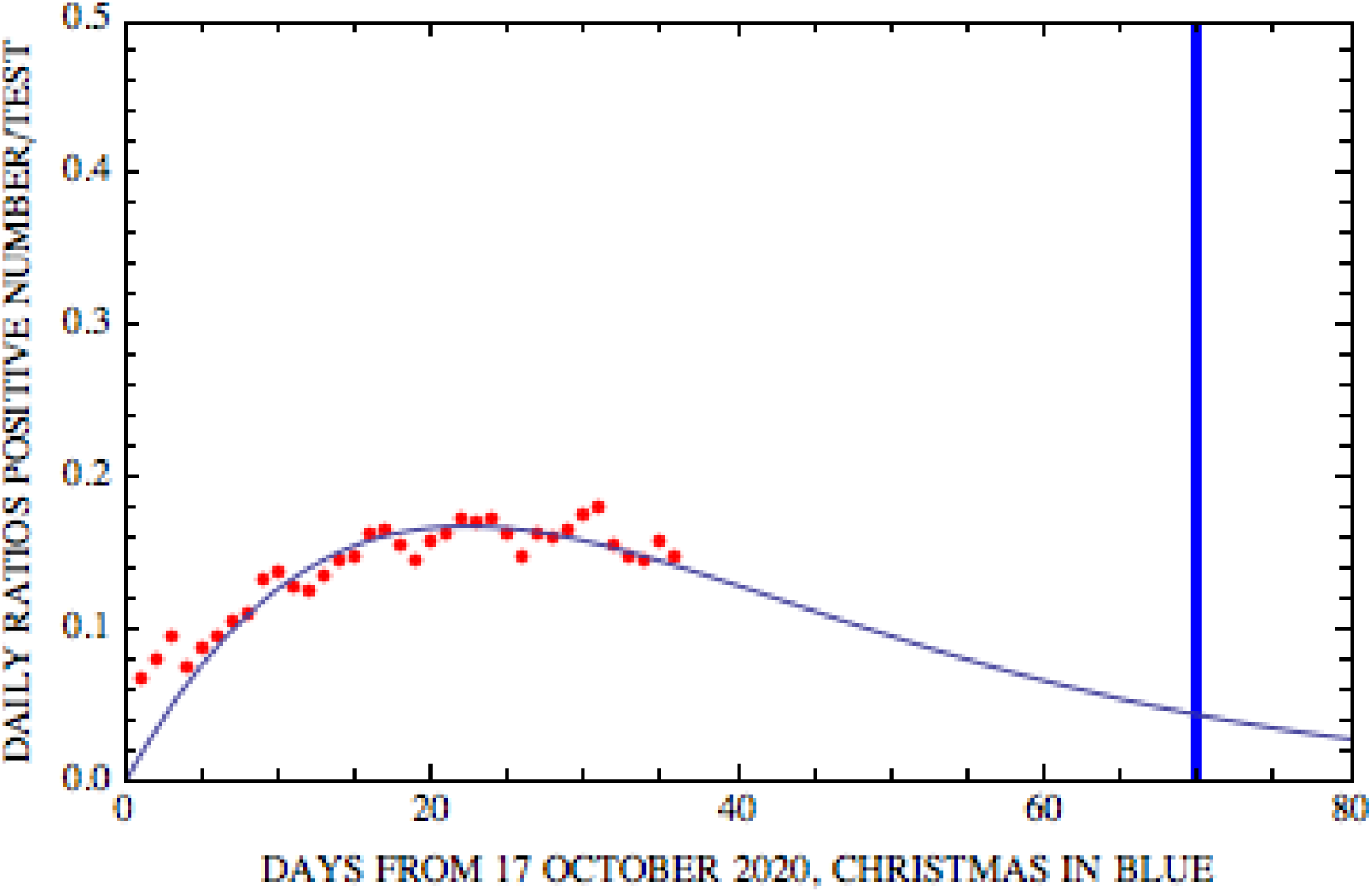
The points in red are the ratios of positive daily cases-to-tests (positivity rates) from 17 October 2020, included, to 21 November 2020, included (36 points). The solid line is the Planck’s law fitting function extrapolated a few weeks after the last data point of 21 November. This figure corresponds to Fig. 4 of ref. [4].

**Fig. 2.**
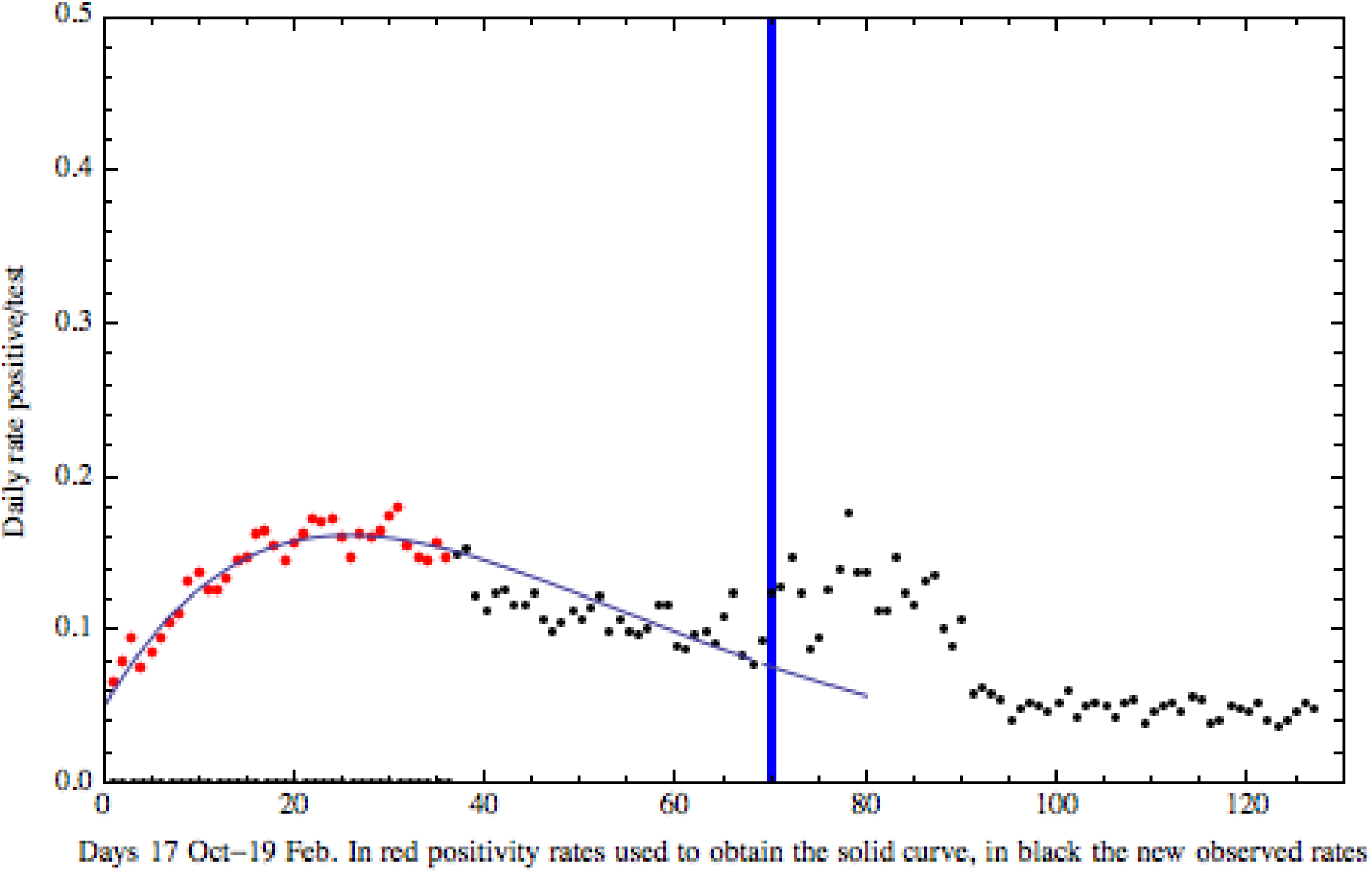
This figure is the same of Fig. 1 where, however, we added, with black circles, the positivity rates (i.e. the ratios of positive daily cases-to-tests) after 21 November 2020, rates not known to us and thus not used to determine the parameters of the Planck’s distribution (solid line). Indeed, the Planck’s distribution fitting function was based on the red points only. The fitting function extrapolated after about one month shows a very good agreement with the observed positivity rates. For example, after 31 days, on 22 December 2020, the difference between the positivity rate predicted by the fitting Planck’s distribution and the real observed positivity rate is less than 2 percent only.

## 2. Prediction of the new wave of pandemic during summer 2021 in Italy

In spite of the diffusion of the Covid vaccines in Italy, probably due to the Covid Delta Variant, starting approximately during the mid of June, it was observed a new wave of pandemic in Italy. In Fig 3, we show the *positivity rates* (i.e., the ratios of positive daily cases-to-tests) from February 26, 2020 until September 12, 2021. In this figure, the new wave of the pandemic in Italy during summer 2021 can be observed after approximately the point 500 of the x-axis.

**Fig. 3.**
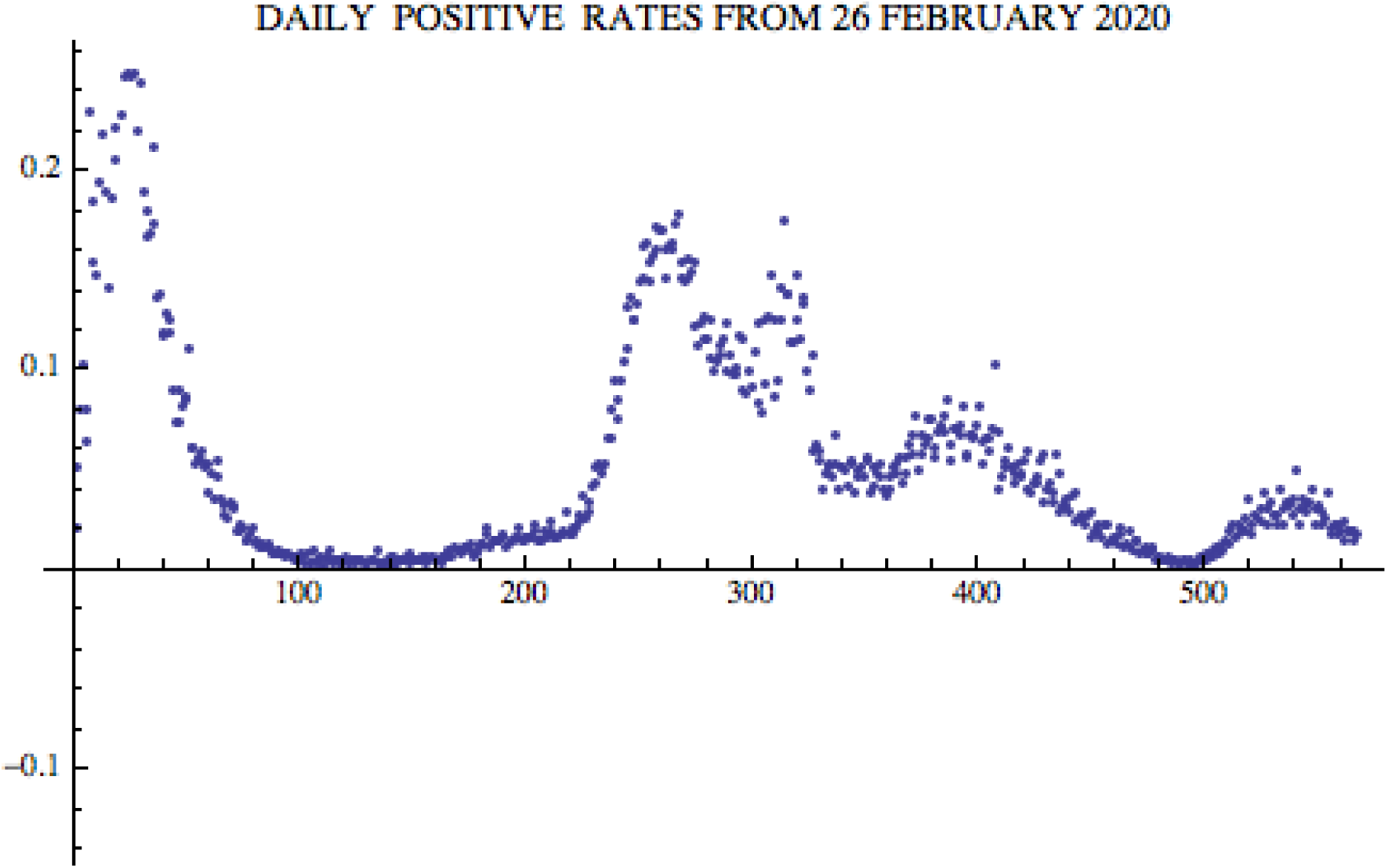
Ratios of positive daily cases-to-tests (*positivity rates*) from February 26, 2020 until September 12, 2021 in Italy. It shows the various waves of the pandemic.

On 9 August 2021, we analyzed and fitted with the Planck’s distribution, with three parameters, the first 30 positivity rates from 10 July 2021 (included) until 8 August 2021 (included). The result is displayed in Fig. 4A, which shows a peak approximately around the middle of August. The agreement of the prediction of this Planck’s function (fitting up to 10 July 2021) and the positivity rate observed after 5 weeks, on 12 September 2021 (the last red point, number 65, of Figs. 4B and 5), is very good. Five weeks later, we then repeated the analysis and the fit using the additional next 35 days of daily positivity rates, i.e., until 12 September 2021 (included). The result is shown in Fig 4B. Even though the daily positivity rates are somehow oscillating, the extended fit of Fig. 4B confirms that the peak of the pandemic occurred approximately around the middle of August 2021.

**Fig. 4A.**
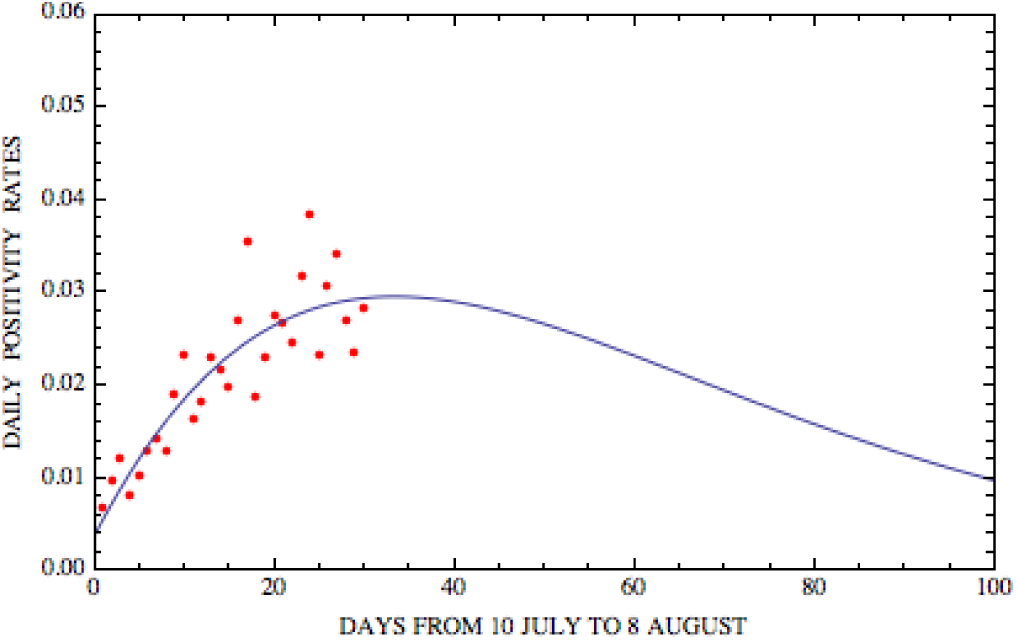
Ratios of positive daily cases-to-tests (daily positivity rates) from 10 July 2021, included, until 8 August 2021, included (30 red points). The solid line is the best fit Planck’s distribution function through the 30 red points. The agreement of the prediction of this Planck’s fitting function and the positivity rate observed after 5 weeks, on 12 September 2021 (the last red point, number 65, of Figs. 4B and 5), is extremely good.

**Fig. 4B.**
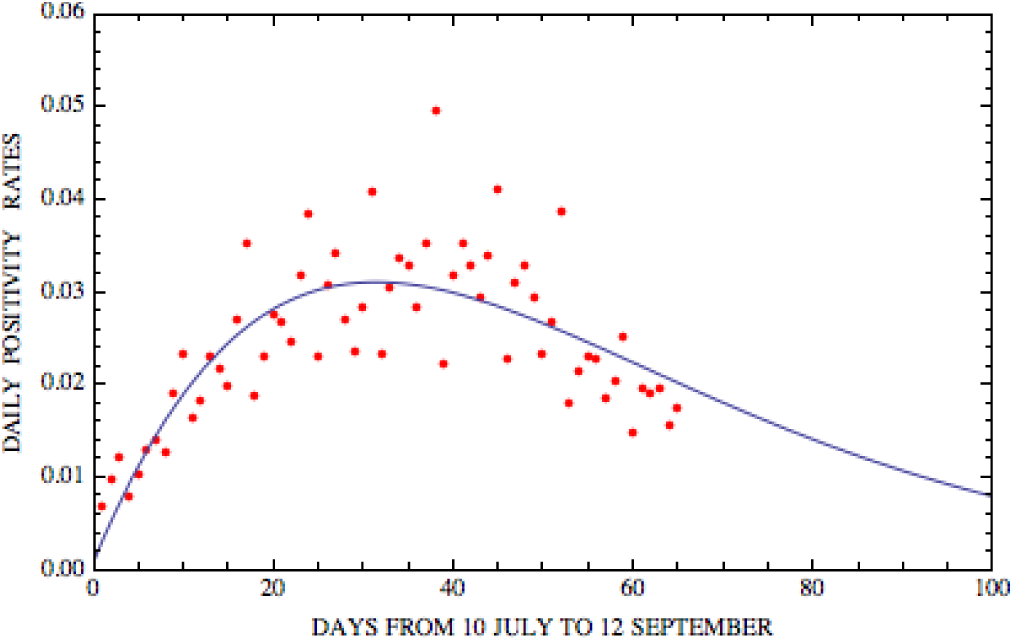
Ratios of positive daily cases-to-tests (daily positivity rates) from 10 July 2021, included, until 12 September 2021 included (65 red points). The solid line is the best fit Planck’s distribution function through the red points.

**Fig. 5.**
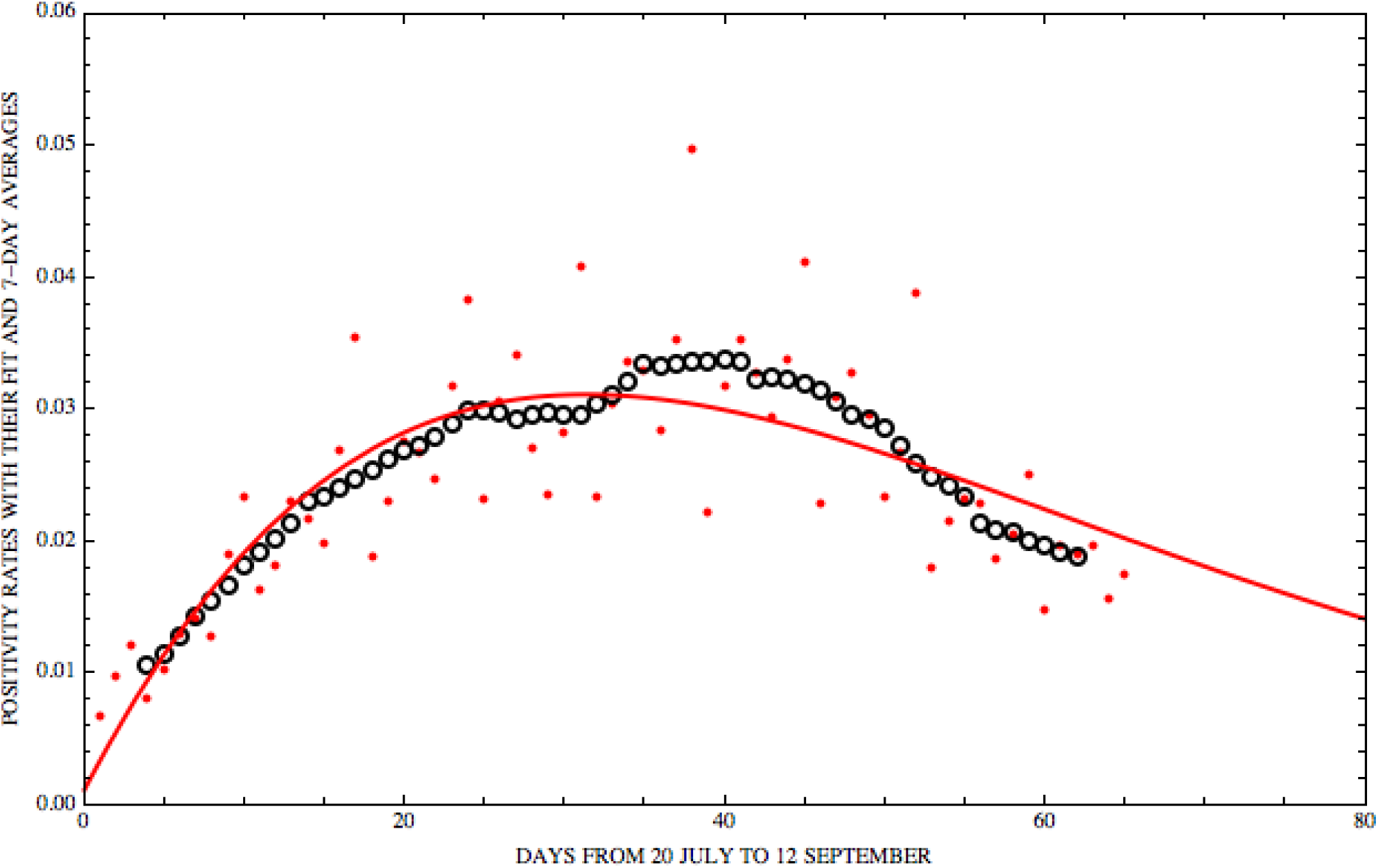
The red points are the ratios of positive daily cases-to-tests (daily positivity rates) from 10 July 2021 until 12 September 2021 and the solid line is the best fit Planck’s function through these red points. The black circles are the 7-day moving averages of the daily positivity rates.

To take into account the oscillating nature of the daily positivity rates during the period middle of July-beginning of September, we took the 7-day moving average of the daily positivity rates. The result is shown in Fig. 5, where the black circles represent the 7-day moving averages of the daily positivity rates, the red points the daily positivity rates and the continuous solid red line is the Planck’s fitting function of the daily positivity rates. Fig. 5 confirms that the peak was reached approximately around the middle of August and that we are now in the decreasing phase of the summer 2021 wave of the pandemic. The end of the summer holidays, the beginning of the activities, schools, intensive use of public transportation, the change in distancing measures and the further diffusion of the vaccines may change again the trend of the positivity rates of the pandemic.

## Conclusions

In a previous paper [1], we carried out an extensive study of a number of distributions to possibly fit the daily positivity rates (ratios of positive daily cases-to-tests) of the Covid-19 pandemic. We concluded that the best fitting function is of the type of the Planck’s law with three parameters. In November 2020, we then applied such fitting function to predict the behavior of the pandemic more than one month later, i.e., during the Christmas holidays 2020, in Italy. Here we show that our prediction, performed in November using the Planck’s distribution, was accurate after over one month, with an error of a few percent only. For example, after 31 days, the difference between our prediction and the observed positivity rate was less than 2%. only We finally apply, the Planck’s distribution, with three parameters, to study the latest wave of the pandemic occurring in Italy during summer 2021. We conclude that the peak of the wave of the pandemic during summer 2021 in Italy was reached approximately around the middle of August and that the prediction of the Plank’s fitting function of the decreasing phase is quite accurate even after 5 weeks after the last datum we used.

## Data Availability

The data of the Covid-19 pandemic were obtained from:
https://github.com/pcm-dpc/COVID-19/blob/master/dati-andamento-nazionale/dpc-covid19-ita-andamento-nazionale.csv

